# Magnetic Resonance Elastography of Regional Aortic Mechanics and True- and False-Lumen Stiffness in Aortic Dissection: A Feasibility Study

**DOI:** 10.64898/2026.09.09.26362667

**Authors:** Adnan A. Hirad, Huiming Dong, Manjunathan Nanjappa, Baqir Kedwai, Kristin Thompson, Joel Kruger, Jarvy Chen, Arunark Kolipaka, Doran Mix

**Affiliations:** Department of Surgery, University of Rochester School of Medicine, Rochester, NY, USA; Department of Neuroscience, University of Rochester School of Medicine, Rochester, NY, USA; Department of Radiation Oncology, University of California San Francisco, Health; Department of Radiology, The Ohio State University, Wexner Medical Center, Columbus, OH, USA

**Keywords:** aorta, aortic dissection, arterial stiffness, magnetic resonance elastography, false lumen, true lumen

## Abstract

**Objective:** Regional aortic mechanics and the mechanical environments of the true and false lumens are not represented by maximal diameter alone. We evaluated whether magnetic resonance elastography (MRE) could provide paired descending thoracic and abdominal aorta stiffness estimates and separate true- and false-lumen estimates in thoracic aortic dissection.

**Methods:** We acquired 60-Hz descending thoracic and abdominal aortic MRE data in 10 healthy subjects and in 2 patients with chronic type B aortic dissections. Three motion-encoding directions and four temporal offsets were reduced to first-harmonic complex wave fields. After directional filtering, we traced a curved centerline through the aortic mask. Local spatial frequency was estimated along arc length and converted to effective shear stiffness. The primary regional outcome was the mean effective stiffness over the central 20%-90% of the path from the quality-ranked slice in each region. In dissection, the whole-aorta mask and flap were defined on a registered sagittal scout; a vascular surgeon assigned the true and false lumens, and separate, manually centered lumen paths followed one union-mask phase correction and unwrapping step. We compared regional values with paired tests.

**Results:** All 10 regional datasets and all 56 available slices were processed without failure. The 10 healthy volunteers (mean age, 26.6 ± 4.6 years; 6 men) had descending thoracic stiffness of 15.43 ± 3.30 kPa and abdominal stiffness of 19.20 ± 3.14 kPa. The mean thoracic-minus-abdominal difference was -3.77 ± 3.40 kPa (95% confidence interval, -6.20 to -1.34 kPa; paired t-test, P=.007; exact Wilcoxon signed-rank test, P=.010). Eight participants had a higher abdominal value, and two had a higher thoracic value. In Subject A with a chronic aortic dissection, slice-averaged true- and false-lumen stiffness was 21.62 and 19.07 kPa, respectively (false-minus-true, -2.54 kPa; absolute percentage difference, 12.5%). In Subject B with a chronic aortic dissection, corresponding values were 12.17 and 17.40 kPa (false-minus-true, +5.22 kPa; 35.3%).

**Conclusions:** We demonstrated the feasibility of MRE to provide paired regional analysis of the aorta in healthy people and to separate stiffness profiles of the true and false lumens in Type B thoracic dissection cases. Abdominal effective stiffness exceeded descending thoracic stiffness in the volunteer cohort. Opposite true- and false-lumen contrast in the two subjects with chronic Tye B aortic dissections demonstrates measurement feasibility but does not establish a consistent compartment pattern, disease threshold, or intrinsic wall material property.

**Clinical Relevance:** Surveillance of aortic disease is primarily anatomic, although expansion and rupture are mechanical processes. A regional MRE measurement may complement anatomic analyses by localizing wave-propagation behavior to a specific aortic segment or dissection lumen. The present study establishes the feasibility of noninvasive measurement of stiffness in a true and false lumen of dissected aorta.

**Highlights:**

- Local-frequency magnetic resonance elastography (MRE) yielded paired abdominal and descending thoracic effective-stiffness estimates in all 10 volunteers.
- Abdominal effective stiffness exceeded descending thoracic effective stiffness in this young feasibility cohort.
- True and false-lumen stiffness contrast was measurable in both dissection cases, but its direction differed between cases.
- The results show the potential of MRE to monitor aortic diseases such as dissection-associated aneurysms and abdominal and thoracic aneurysms.

## Introduction

Aneurysmal aortic disease is assessed principally by anatomical features such as maximal diameter, interval growth, branch-vessel involvement, rupture-related findings, and, in dissection, true and false-lumen morphology and patency. These variables remain central to clinical decision-making (*1*). However, these anatomic features do not describe the vessel’s mechanical state, which likely contributes to further deterioration and rupture. Aortic stiffness depends on extracellular matrix architecture, smooth muscle tone, wall geometry, prestress, and blood pressure. Clinically, large-artery stiffness measured by pulse-wave velocity has been shown to be associated with cardiovascular events and mortality (*2, 3*).

The aorta is mechanically heterogeneous. Thoracic and abdominal segments differ in elastin content, collagen organization, wall thickness, curvature, branch loading, and age-related remodeling (*4–6*). Ex vivo measurements demonstrate longitudinal variation in human aortic structure and biaxial response (*4*), whereas in vivo pulse-wave measurements show region-specific aging effects (*5, 6*). As such, a global stiffness metric cannot determine whether a change is concentrated in the thoracic or abdominal aorta. Regional measurement is therefore critical to elucidating segmental pathologic differences.

In type B aortic dissections, persistent false-lumen degeneration contributes to late aneurysmal enlargement and reintervention, necessitating lifelong cross-sectional imaging (*7–9*). Although false-lumen patency and hemodynamics are associated with adverse remodeling, conventional measures of morphology and flow do not directly characterize local mechanical properties (*9–12*). Separate assessment of true- and false-lumen stiffness could therefore provide a complementary mechanical marker for monitoring dissection remodeling and disease progression.

Magnetic resonance elastography (MRE) is a noninvasive technique to estimate the stiffness of soft tissues by synchronizing external vibrations with the motion encoding gradients in the phase of MR images(*13–16*). Aortic MRE studies have demonstrated feasibility to estimate stiffness in abdominal aortas, pressure sensitivity, associations with age and pulse-wave velocity, cardiac-cycle dependence, and measurement in abdominal aortic aneurysm (*17–25*). Multifrequency MRE has also measured ascending thoracic, descending thoracic, and abdominal aortic segments with excellent reader agreement and reproducibility; in 20 healthy participants, descending thoracic and abdominal wave speeds were similar (*17*). Our prior feasibility study demonstrated single-breath-hold thoracic aortic MRE and stiffness contrast in two-lumen dissection phantoms (*26*).

This study had two aims: (1) to assess the feasibility of aortic MRE for paired descending thoracic and abdominal effective-stiffness measurements in volunteers, and (2) to demonstrate separate true- and false-lumen effective-stiffness measurements in two cases of thoracic aortic dissection.

## Methods

### Study Design and Participants

The study was conducted after the approval of institutional review board and a written informed consent was obtained. The regional cohort comprised 10 volunteers with both abdominal and descending thoracic examinations (age range, 20-32 years; mean, 26.6 ± 4.6 years; 6 male and 4 female) and two patients with chronic residual type B aortic dissections: subject A (female, 78 years) and subject B (male, 70 years) (Table I). The present analysis used no demographic or clinical covariates beyond age and sex.

**Table I.** Cohort characteristics.

| Cohort Characteristics |  |  |
| --- | --- | --- |
| Regional cohort | Participants | 10 with paired abdominal and thoracic MRE |
|  | Age | 26.6 ± 4.6 years (range, 20-32) |
|  | Sex | 6 (male); 4 female |
| Dissection cases | Subject A | Male, 78 years; thoracic |
| Dissection cases | Subject B | Female, 70 years; thoracic |

### MRE Acquisition

All imaging was performed on a 3-T MAGNETOM Prisma Fit system (Siemens Healthineers, Erlangen, Germany) using a flow-compensated spin-echo echo-planar MRE sequence during a single-breath hold. External vibarations of 60Hz was applied both to the thoracic and abdominal region. Each acquisition contained four equally spaced temporal offsets and three orthogonal motion-encoding directions. The in-plane field of view was 300 x 300 mm, the reconstructed matrix was 192 x 192 pixels, the reconstructed pixel spacing was 1.5625 x 1.5625 mm, and nominal slice thickness was 10 mm. Table II summarizes the acquisition details.

**Table II.**
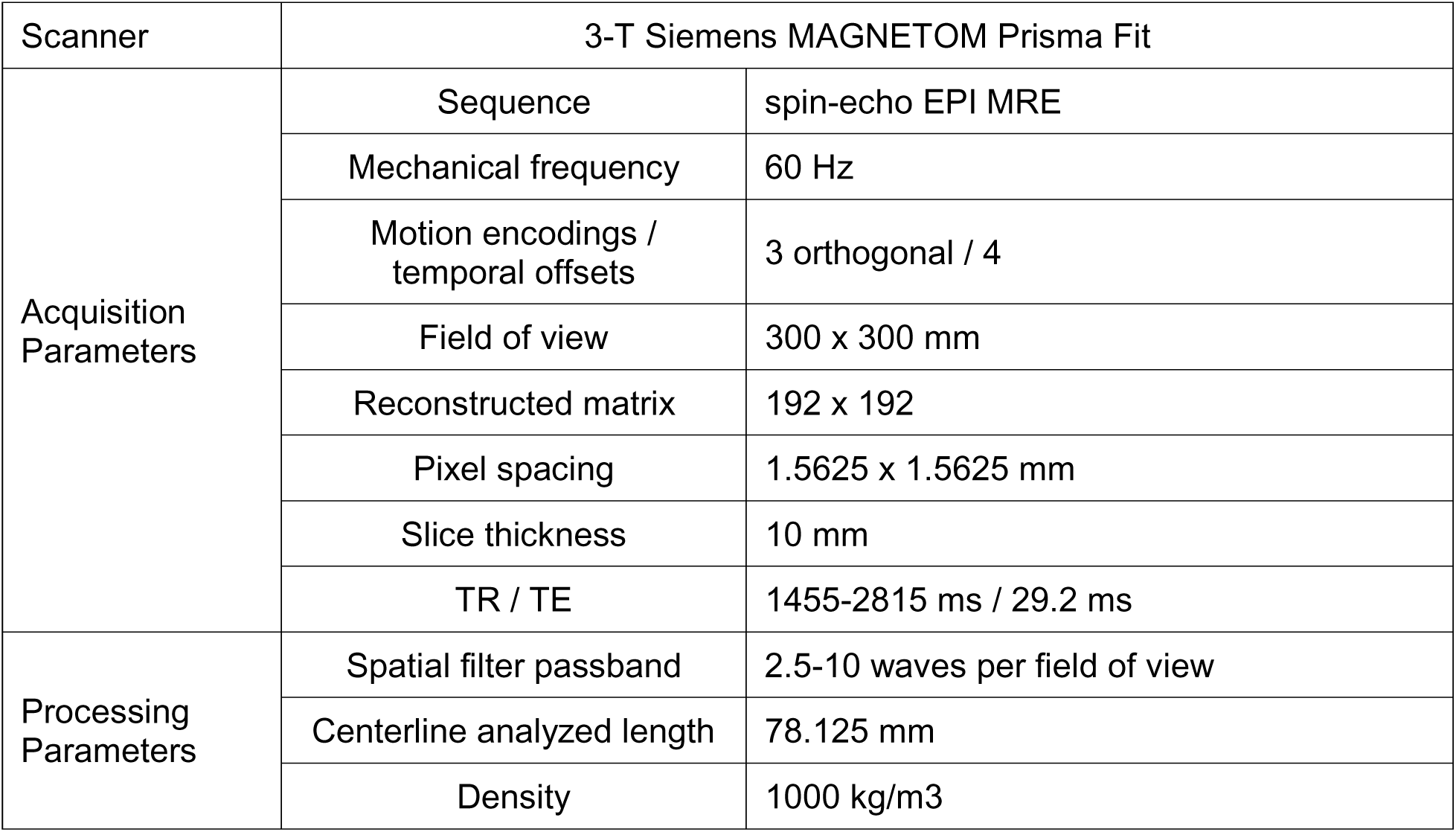
Scan and Acquisition Parameters.

**Table III.**
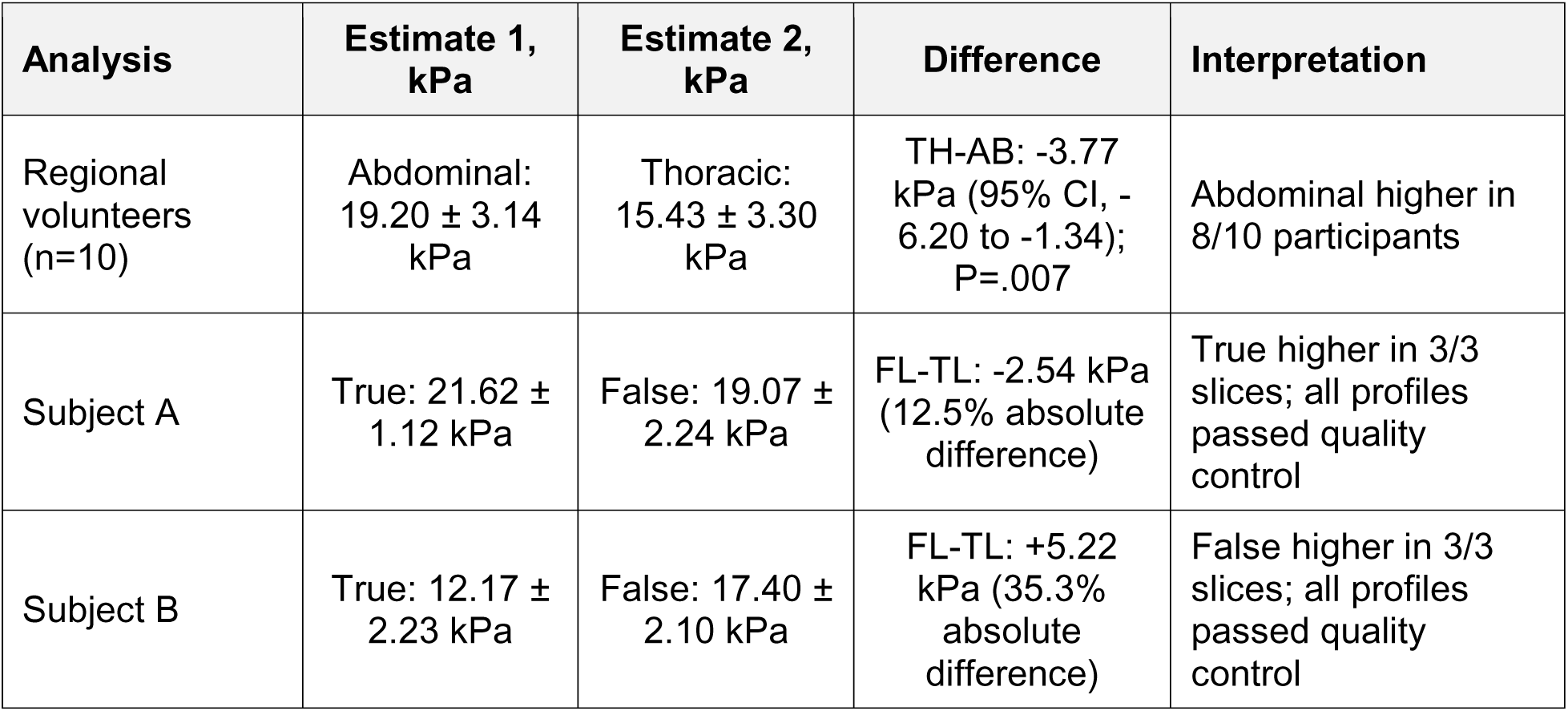
Primary regional and dissection-compartment results.

### Aortic Region and Dissection Lumen Selection

The descending thoracic and infrarenal abdominal aorta were delineated using the registered sagittal anatomic scout together with the corresponding MRE magnitude images. The thoracic region of interest began distal to the aortic arch and extended caudally within the descending thoracic aorta while remaining proximal to the diaphragm. The abdominal region of interest began immediately distal to the level of the renal artery origins and extended caudally toward, but not beyond, the aortic bifurcation. The sagittal scout provided the clearest anatomic landmarks, while the matched MRE magnitude image confirmed that the mask enclosed the intended aortic segment in the MRE acquisition plane. A smooth centerline was generated within each accepted aortic mask and sampled at uniform intervals according to arc length. Arc length measures distance along the curved vessel rather than the shorter straight-line distance between two image coordinates, thereby providing an anatomically appropriate spatial axis for wavelength estimation. A 78.125-mm centerline segment, corresponding to approximately 8 cm. The same physical sampling length was used in the thoracic and abdominal regions to support a paired regional comparison. The analysis workflow is illustrated in Figure 1.

**Figure 1.**
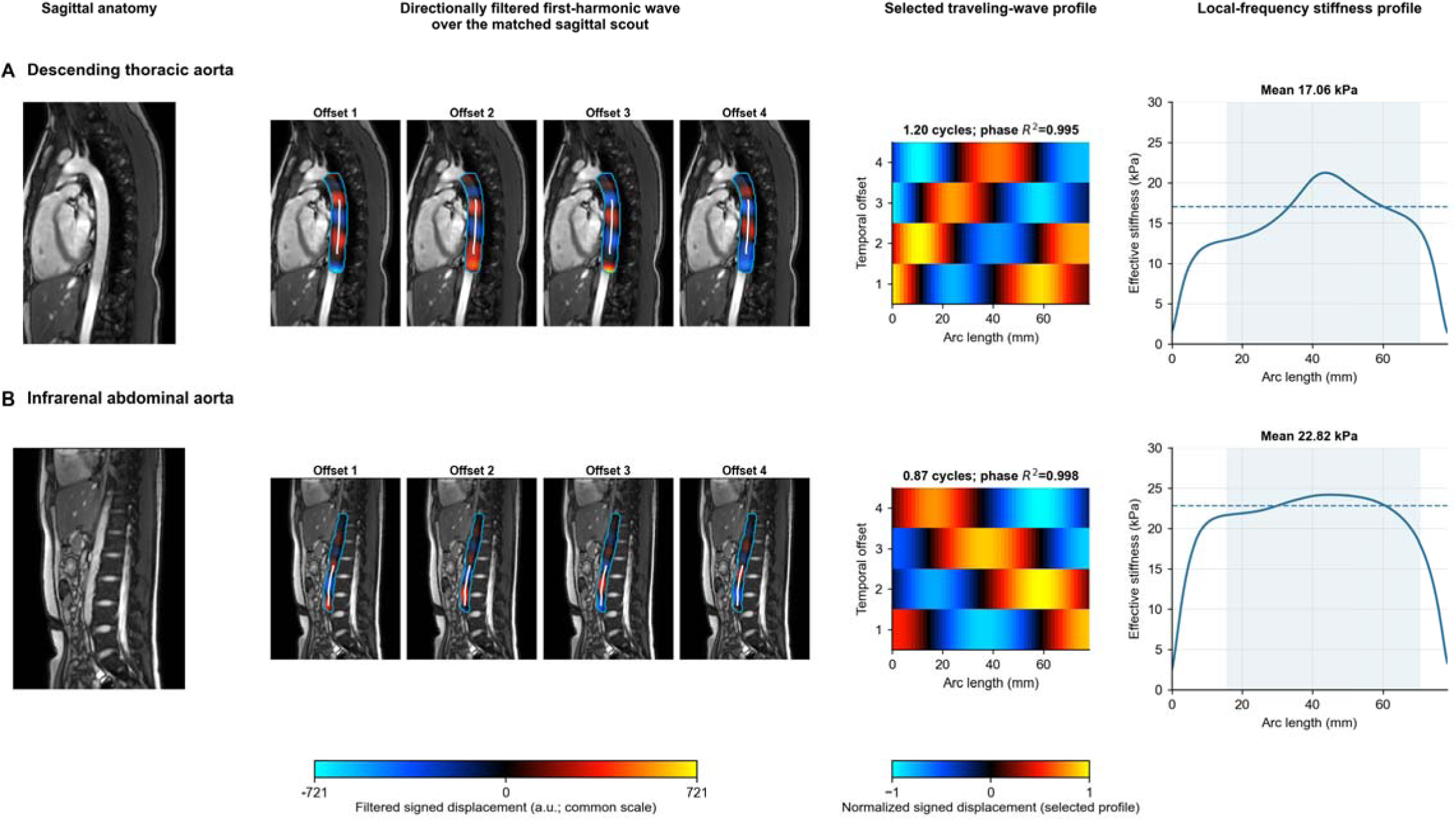
Representative regional aortic MRE measurements. (A) Infrarenal abdominal and (B) descending thoracic measurements from participant C0001. Columns show sagittal anatomy, directionally filtered four temporal wave offsets superimposed on the matched sagittal scout, the selected traveling-wave profile sampled along the curved path, and the local-frequency stiffness profile. Cyan outlines indicate the aortic masks, and white lines indicate the selected sampling paths. Temporal-offset images are displayed without amplitude normalization using a common arbitrary-unit scale across regions. Selected traveling-wave profiles are independently normalized to their maximum absolute amplitude and the colors show phase progression rather than absolute motion amplitude. The shaded stiffness-profile interval marks the central 20%–90% used for numerical summary. Central-path estimates abdominal and the thoracic regions were 22.82 kPa and 17.06 kPa respectively.

For dissection cases, the complete thoracic aortic mask was drawn on a DICOM-registered sagittal scout matched to each MRE slice, and the visible flap was traced within that mask. The flap divided the mask into two compartments. A vascular surgeon reviewed the scout and matched MRE magnitude images and assigned the true and the false lumens in both cases. These assignments were fixed before wave inversion and final compartment summaries.

### Image Processing and Stiffness Estimation

All processing was performed in MATLAB R2024a (MathWorks, Natick, Mass) using custom code. Wrapped phase images were unwrapped before the four temporal offsets for each motion-encoding direction were transformed to the complex first harmonic. For regional analysis, the aortic mask was reduced to a smooth centerline, ordered along its principal axis, and resampled by arc length; when available, an analysis path of 78.125 mm was used. For dissection, phase unwrapping was performed once within the whole-aorta union mask. A common whole-aorta propagation direction was then applied to both lumens on a given slice, while the true- and false-lumen sampling paths were separately centered within their accepted compartments. This kept preprocessing and directional filtering common while allowing each measurement path to follow its lumen.

The complex field for each encoding was directionally filtered over 2.5-10 waves per field of view using a fourth-order butterworth bandpass filter. We analyzed opposing propagation directions separately to reduce cancellation from counter-propagating waves (9). We interpolated the filtered complex wave along centerline arc length (s). For an approximately traveling harmonic wave,

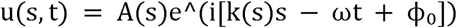

where A(s) is local amplitude, k(s) is local wave number, ω=2πf is angular frequency, f=60 Hz, and □_₀_ is phase offset. For the local-frequency estimation we used an 11-band log-Gaussian spatial-frequency filter bank to estimate wavelength from spatial frequency. The effective stiffness for each component was calculated from wave speed under the conventional local homogeneous, isotropic, linear wave approximation:

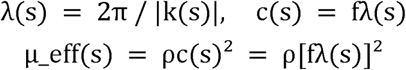

with assumed density ρ=1000 kg/m³. This quantity is reported as effective stiffness because the aortic lumen-wall system is pressurized, curved, thin-walled, anisotropic, and wave-guiding. As such, the simplified conversion is not intended to serve as an intrinsic wall shear modulus.

The six component estimates (three motion encoding diections x two afforementioned propagation directions) were combined using first-harmonic-amplitude-squared weights:

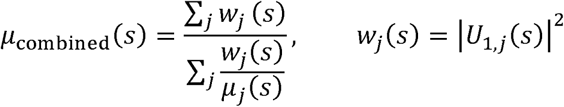

Where:

*s*: is the arc-length position along the accepted curved aortic path, measured in millimeters.
*j*: represents both the motion-encoding component and propagation direction, giving up to six estimates: X-forward, X-reverse, Y-forward, Y-reverse, Z-forward, and Z-reverse.
*μ_j_*(s): is the local effective stiffness obtained by applying 1D local-frequency estimation to component *j*, in kPa
*U*_1,*j*_(s): is the complex first temporal harmonic of directionally filtered component *j*, sampled along the accepted path:

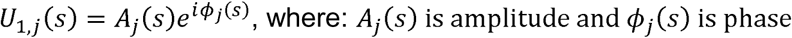

The magnitude of the complex first harmonic, is its amplitude:

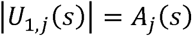

The weight assigned to each component is therefore:

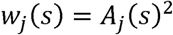

All together, a direction with a stronger measured wave receives more influence than a direction with weak signal(i.e, the amplitude-weighted harmonic mean gives greater influence to components with measurable first-harmonic motion while limiting domination by very large local stiffness values). To reduce endpoint artifacts introduced by path interpolation and the finite spatial support of the local-frequency filters, the first 20% and final 10% of the sampled path were excluded from the primary stiffness summary.

### Quality Control and Slice Selection

Traveling-wave coherence was assessed using the motion-encoding and propagation-direction combination with the greatest mean first-harmonic amplitude. Complex first-harmonic phase was unwrapped along centerline arc length and fitted to a constant-wavenumber model, φ(s)=ks+φ₀, using amplitude-squared-weighted least-squares regression. The resulting weighted coefficient of determination (phase R²) quantified the proportion of amplitude-weighted phase variation explained by steady phase progression along the selected path. Values approaching 1 were considered more consistent with a dominant traveling-wave component, whereas lower values indicated departures from a single linear phase progression. Phase R² was used as a descriptive quality measure and was not entered into the calculation of the primary local-frequency stiffness estimate. Observed cycles were calculated as

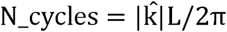

where k is the fitted phase slope and L is analyzed path length. A given slice was included if it had at least 0.5 observed cycles, phase R²≥0.50, and adequate path length. For each volunteer region, passing slices were ranked by the product of dominant first-harmonic amplitude and phase R².

### Statistical Analysis

Continuous results are reported as mean ± standard deviation unless otherwise stated. Descending thoracic and abdominal aorta primary values were compared within participants by paired *t* test, with a two-sided 95% confidence interval for the mean difference. The exact Wilcoxon signed-rank test was used as a nonparametric sensitivity analysis. No multiplicity correction was required because there was one prespecified regional comparison. Dissection-compartment results are reported descriptively as slice-averaged values and false-minus-true differences. With only two cases, no inferential test was performed for subjects with chronic type B dissections. Analyses were conducted in Python 3.11 with SciPy and two-sided *P*<.05 was considered statistically significant.

## Results

### Processing Feasibility and Wave Quality

All 10 volunteer regional datasets and 54 out 56 available slices passed the prespecified wave-quality control criteria. The selected regional measurements all passed quality control and had a median dominant phase R² of 0.985 (range, 0.803-1.000) and a median of 0.90 observed cycles (range, 0.55-1.31). Fifteen of 20 selected regional measurements contained fewer than one complete cycle, but all exceeded the prespecified 0.5-cycle threshold. Representative descending thoracic and abdominal analyses are shown in Figure 1. Complete slice-level measurements are shown in Supplemental Figure 1.

### Paired Abdominal and Descending Thoracic Stiffness

Descending thoracic aorta effective stiffness was 15.43 ± 3.30 kPa (range, 10.42-19.56 kPa) and abdominal aortic effective stiffness was 19.20 ± 3.14 kPa (range, 15.08-24.39 kPa). The paired thoracic-minus-abdominal difference in stiffness was -3.77 ± 3.40 kPa (95% confidence interval, -6.20 to -1.34 kPa; paired t-test, P=.007). The exact Wilcoxon signed-rank test also supported a regional difference (P=.010). Eight participants had a higher abdominal stiffness, and two had a higher thoracic stiffness (Figure 2). Participant-level values are provided in Supplemental Table I.

**Figure 2.**
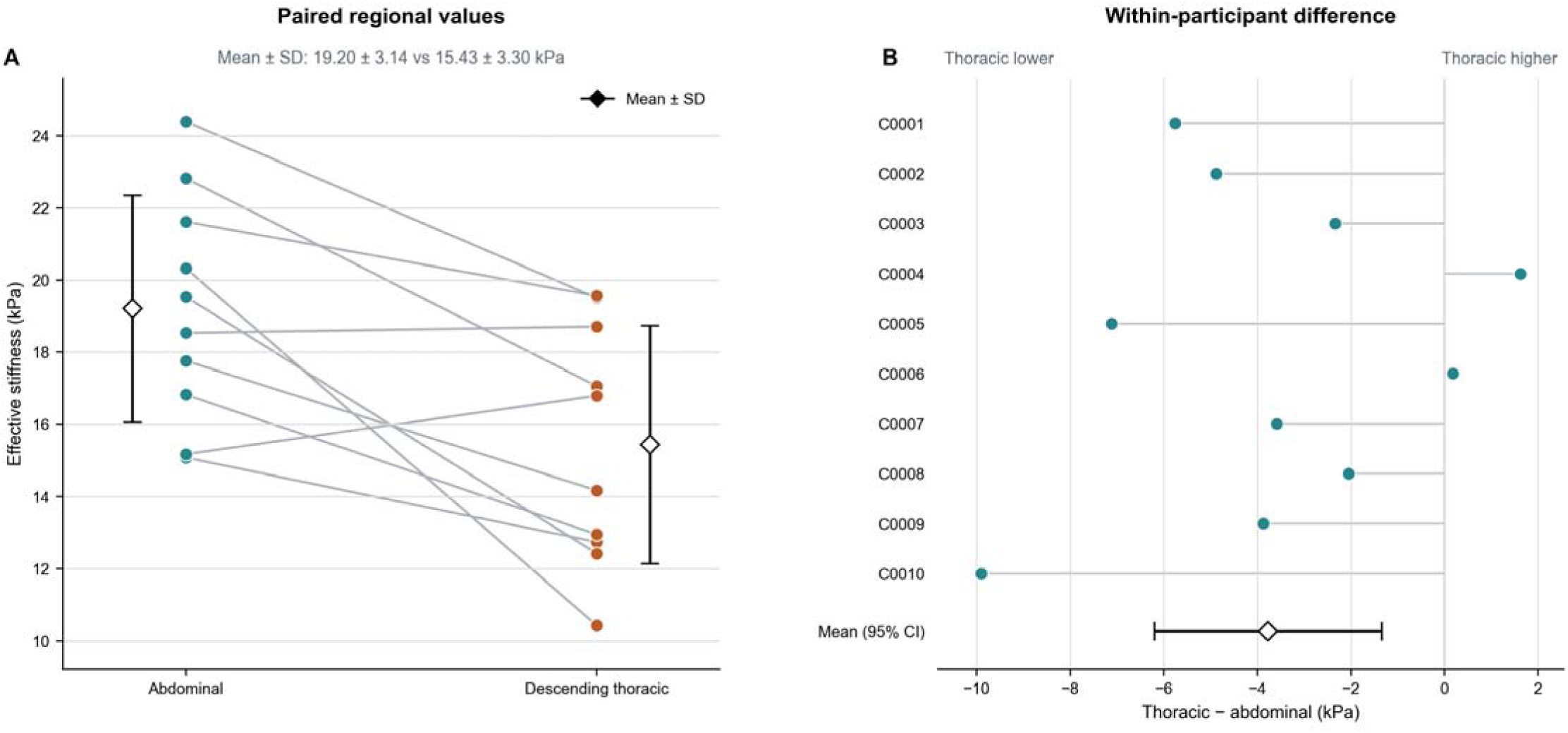
Paired abdominal and descending thoracic effective stiffness. Lines connect the abdominal and descending thoracic primary estimates for each of 10 participants. Black diamonds and error bars show group means and 95% confidence intervals. The thoracic-minus-abdominal stiffness mean difference was -3.77 kPa (95% confidence interval, -6.20 to -1.34 kPa; paired t test, P=.007). Eight paired differences were negative and two were positive.

### Thoracic True- and False-Lumen Stiffness

Both chronic type B thoracic dissections had three analyzable slices. In subject A, slice-specific true-lumen stiffness values were 22.50, 22.00, and 20.35 kPa; false-lumen stiffness values were 21.61, 17.36, and 18.25 kPa. Slice-averaged stiffness was 21.62 ± 1.12 kPa in the true lumen and 19.07 ± 2.24 kPa in the false lumen, a false-minus-true lumen stiffness difference was -2.54 kPa (absolute percentage difference, 12.5%). True-lumen stiffness was higher in all three slices. Phase R² ranged from 0.960 to 0.995, observed cycles ranged from 0.62 to 0.95, and all six lumen-specific measurements passed the prespecified quality control criteria.

Subject B, slice-specific true-lumen stiffness values were 10.49, 11.33, and 14.70 kPa; false-lumen stiffness values were 15.07, 19.14, and 17.98 kPa. Slice-averaged stiffness was 12.17 ± 2.23 kPa in the true lumen and 17.40 ± 2.10 kPa in the false lumen, a false-minus-true lumen stiffness difference was +5.22 kPa (35.3%). False-lumen stiffness was higher in all three slices. Phase R² ranged from 0.884 to 0.999, observed cycles ranged from 0.54 to 1.34, and all six lumen-specific measurements passed quality control. Figure 3 shows representative compartment results, and complete slice-level quality metrics are shown in Supplemental Figures 2 and 3. Based on clinical imagig data, maximum aortic diameter was greater in Subject B than in Subject A (∼ 43 vs 38 mm). Conversely, the dissection was more extensive in Subject A, extending from aortic zone 3 through zone 10, compared with zones 3 through 5 in Subject B.

**Figure 3.**
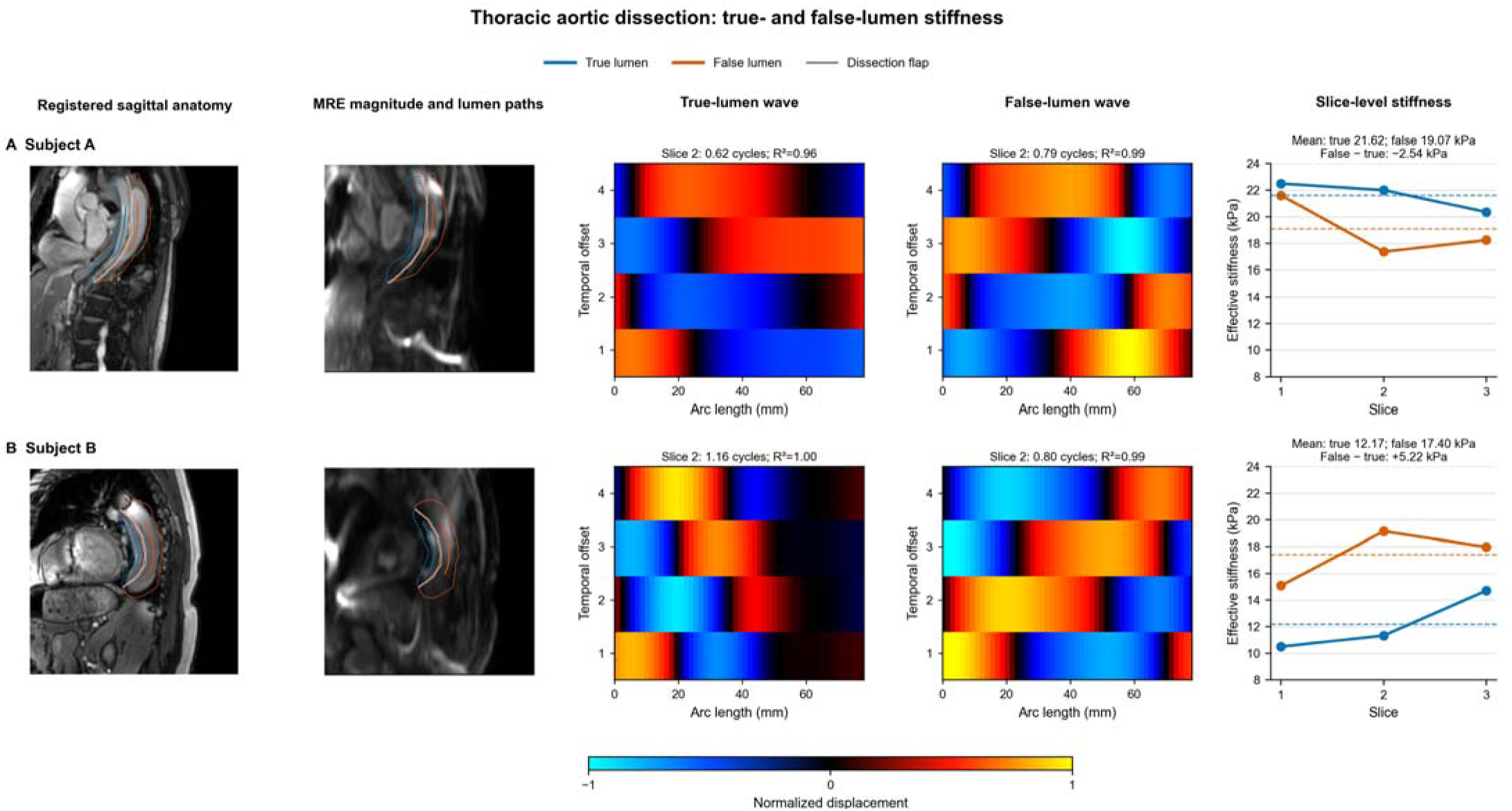
Thoracic true- and false-lumen stiffness in two dissection cases. Columns show sagittal anatomy, matched MRE magnitude with vascular-surgeon-assigned true-lumen (blue) and false-lumen (orange) paths, directionally filtered true- and false-lumen traveling-wave profiles from the middle slice, and all slice-level effective-stiffness values. Wave color encodes signed displacement normalized within each displayed lumen. Dashed lines indicate slice-averaged lumen values. True-lumen stiffness exceeded false-lumen stiffness in all three slices for Subject A, whereas false-lumen stiffness exceeded true-lumen stiffness in all three slices for Subject B. All 12 lumen-specific measurements passed the prespecified quality control criteria.

## Discussion

In this feasibility study, MRE enabled paired estimates of effective stiffness in the descending thoracic and abdominal aorta. Among 10 healthy volunteers, abdominal effective stiffness exceeded descending thoracic stiffness by 3.77 kPa on average; the 95% confidence interval excluded zero, and 8 of 10 participants showed the same direction. We further applied the approach to the true- and false-lumens in two cases of type B aortic dissection. True-lumen effective stiffness was higher in all three slices of subject A, whereas false-lumen effective stiffness was higher in all three slices of subject B. Together, these findings support the feasibility of regional and compartment-specific aortic MRE while showing that lumen contrast was case dependent in this small series.

The thoracic-abdominal difference observed here contrasts with the similar descending thoracic and abdominal shear-wave speeds reported by Schaafs et al using steady-state multifrequency MRE (*17*). The studies differ in drive-frequency sampling, acquisition, inversion output, and cohort; as such, their absolute measurements are not directly interchangeable. Regional aortic structure and mechanics also vary with composition, geometry, pressure, and age (*4–6*). Therefore, while our result demonstrates detectable paired regional contrast, larger multi-site sample sizes are needed to establish reference values and their expected direction across age and disease.

The effective-stiffness estimates in this study were higher than values reported with some multifrequency wave-speed approaches. MRE-derived stiffness values depend on drive frequency, acquisition timing, directional filtering, spatial resolution, and inversion model (*16–18, 25*). We obtained our estimates at 60 Hz using 1D local-frequency analysis and a density-based wave relation. They should therefore be interpreted within this acquisition and analysis context and are not directly interchangeable with wave speed, Young’s modulus, distensibility, or constitutive wall parameters. Until cross-method calibration is available, within-participant and within-protocol comparisons are likely to be more informative than direct comparison of absolute values across MRE techniques.

The true- and false-lumens are exposed to different flow patterns, pressure histories, boundary conditions, thrombus burdens, and degrees of flap and wall support. The presence of a measurable contrast in both cases is therefore plausible, but its opposite direction in subject A and subject B argues against assigning a universally stiffer lumen from this series. Persistent false-lumen patency is associated with adverse remodeling and late aortic events (*9*). In addition, 4D-flow MRI studies have linked false-lumen ejection and relative-pressure measures with subsequent aortic growth, while tear location and size can influence false-lumen pressure (*10–12*). Compartment-specific effective stiffness may complement anatomic and flow-based measurements by characterizing another aspect of the mechanical environment within a dissected aorta.

The measured response represents the behavior of a coupled system that includes blood transmitting mechanical motion, the intimal flap, residual media and adventitia, surrounding tissue, and in vivo prestress. The true- and false-lumen contrast should therefore be interpreted as a compartment-specific effective mechanical phenotype rather than as an isolated measurement of wall material properties. Its clinical value will depend on reproducibility and on whether longitudinal changes correspond to false-lumen thrombosis, aneurysmal enlargement, or other features of dissection remodeling. Prospective studies should combine MRE with blood pressure, cardiac phase, contrast-enhanced anatomic imaging, false-lumen patency and thrombus characterization, tear morphology, and longitudinal measurements of aortic growth.

The curvilinear sampling approach permits the analysis coordinate to follow the vessel rather than intersecting a curved aortic segment with a straight line. Arc length consequently provides an anatomically meaningful distance axis for wavenumber estimation. In dissection cases, applying a common whole-aorta filtering direction while sampling the true and false lumens separately also allows compartment comparisons without independently optimizing the directional filter for each lumen. These features support consistent regional and compartment-level analysis while retaining the established directional-filtering and local-frequency operations used in MRE (*15–18*).

## Limitations

This study was small, and the volunteer cohort was young and had a narrow age range; therefore, the findings cannot establish reference intervals, age or sex effects, or disease thresholds. The dissection analysis included only two cases. Although each case showed a consistent within-case direction across slices, the direction differed between cases. These findings are hypothesis-generating and do not establish prevalence, diagnostic performance, prognosis, or treatment response.

The acquisition was performed at a single mechanical frequency, and effective stiffness was calculated using a simplified density-based wave relation. The aorta is anisotropic, viscoelastic, prestressed, thin-walled, and surrounded by heterogeneous tissues. The reported values therefore reflect effective stiffness under the study conditions rather than intrinsic material modulus.

Spatial wave coverage was limited in some measurements. Fifteen of 20 selected regional paths and several dissection-lumen paths contained fewer than one complete wave cycle, although all selected regional and lumen-specific measurements exceeded the 0.5-cycle threshold. Although local-frequency estimation can operate near the spatial sampling limit (*18*), longer coherent wave trains would increase confidence in the resulting estimates. To provide transparent indicators of this limitation, we calculated amplitude-weighted phase-fit R² and observed-cycle measurements. We ranked slice selection by quality rather than pooling by volume, and one abdominal dataset contained only one available slice.

## Conclusions

Overall, this study demonstrated the feasibility of using MRE for paired regional aortic stiffness measurements and separate true and false-lumen analysis in dissection. Abdominal effective stiffness exceeded descending thoracic stiffness in this volunteer cohort, while both dissection cases showed lumen contrast stiffness with opposite directions. Our proposed approach could be used in prospective studies of regional aortic mechanics and dissection remodeling.

## Data Availability

All data produced in the present study are available upon reasonable request to the authors

## Declarations

### Ethics Approval and Consent to Participate

### Data and Code Availability

All data required for interpreting results are in the data. Code used to analyze the data could be shared upon reasonable request.

---

**Supplemental Table I.**
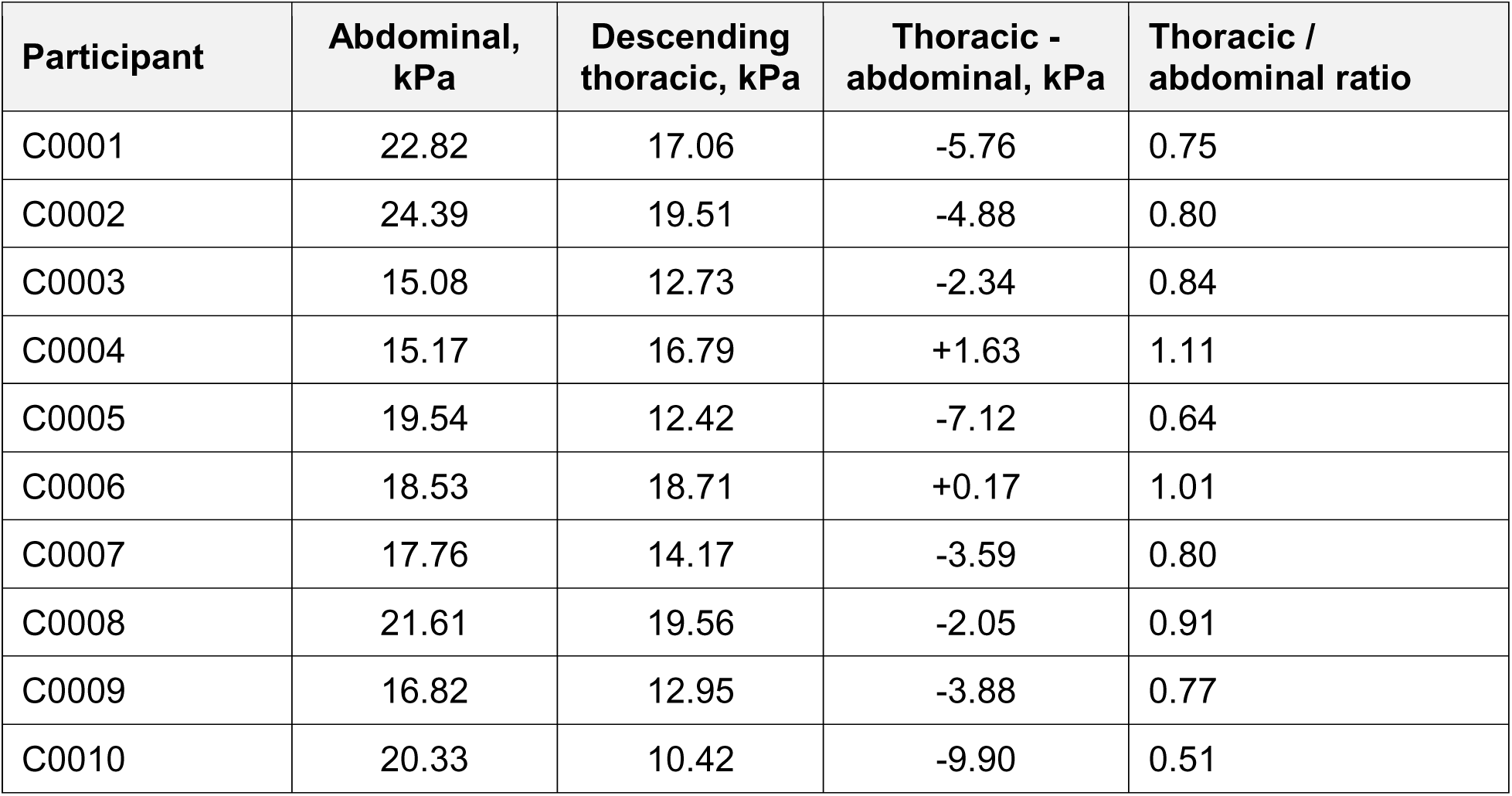
Individual paired regional values.

| Participant | Abdominal, kPa | Descending thoracic, kPa | Thoracic - abdominal, kPa | Thoracic / abdominal ratio |
| --- | --- | --- | --- | --- |
| C0001 | 22.82 | 17.06 | -5.76 | 0.75 |
| C0002 | 24.39 | 19.51 | -4.88 | 0.80 |
| C0003 | 15.08 | 12.73 | -2.34 | 0.84 |
| C0004 | 15.17 | 16.79 | +1.63 | 1.11 |
| C0005 | 19.54 | 12.42 | -7.12 | 0.64 |
| C0006 | 18.53 | 18.71 | +0.17 | 1.01 |
| C0007 | 17.76 | 14.17 | -3.59 | 0.80 |
| C0008 | 21.61 | 19.56 | -2.05 | 0.91 |
| C0009 | 16.82 | 12.95 | -3.88 | 0.77 |
| C0010 | 20.33 | 10.42 | -9.90 | 0.51 |

## Supplemental Figure Legends

**Supplemental Figure 1.**
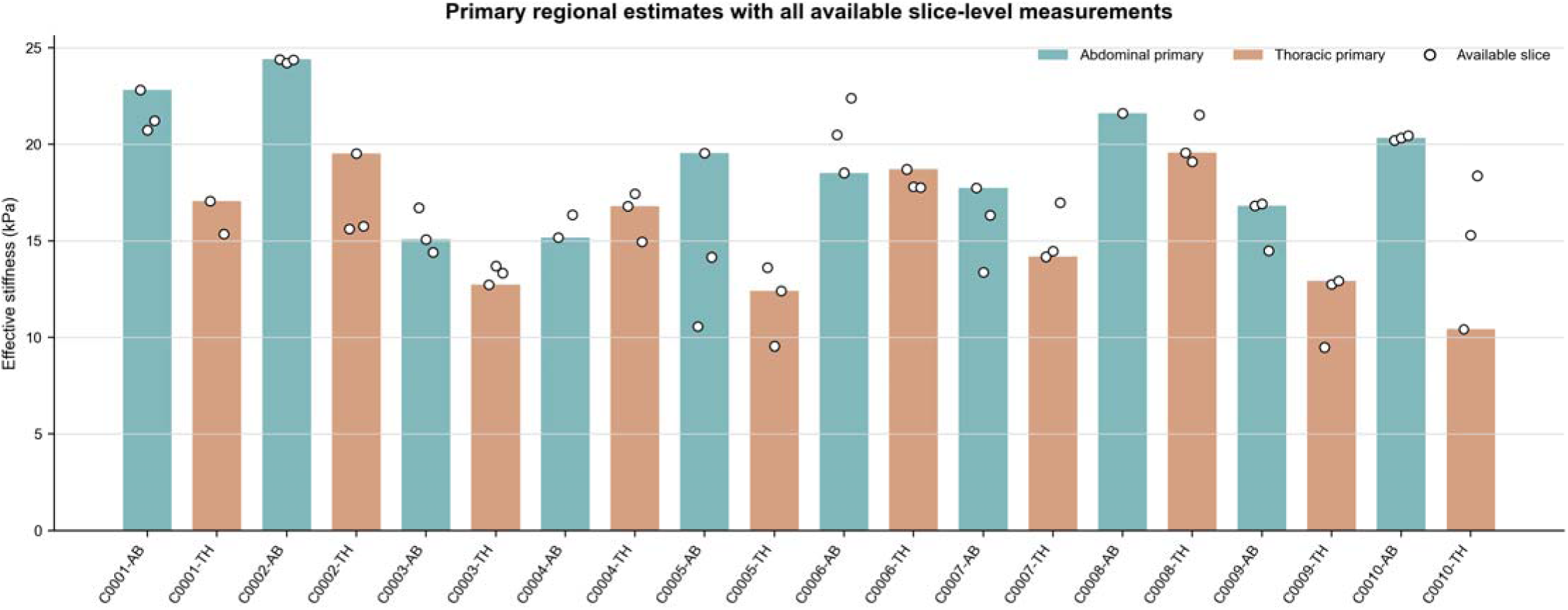
Regional slice-level quality review. Bars show the automatically selected primary slice for each participant and region. White points show all 56 analyzed slice values. Slice values are repeated technical measurements and were not treated as independent participants.

**Supplemental Figure 2.**
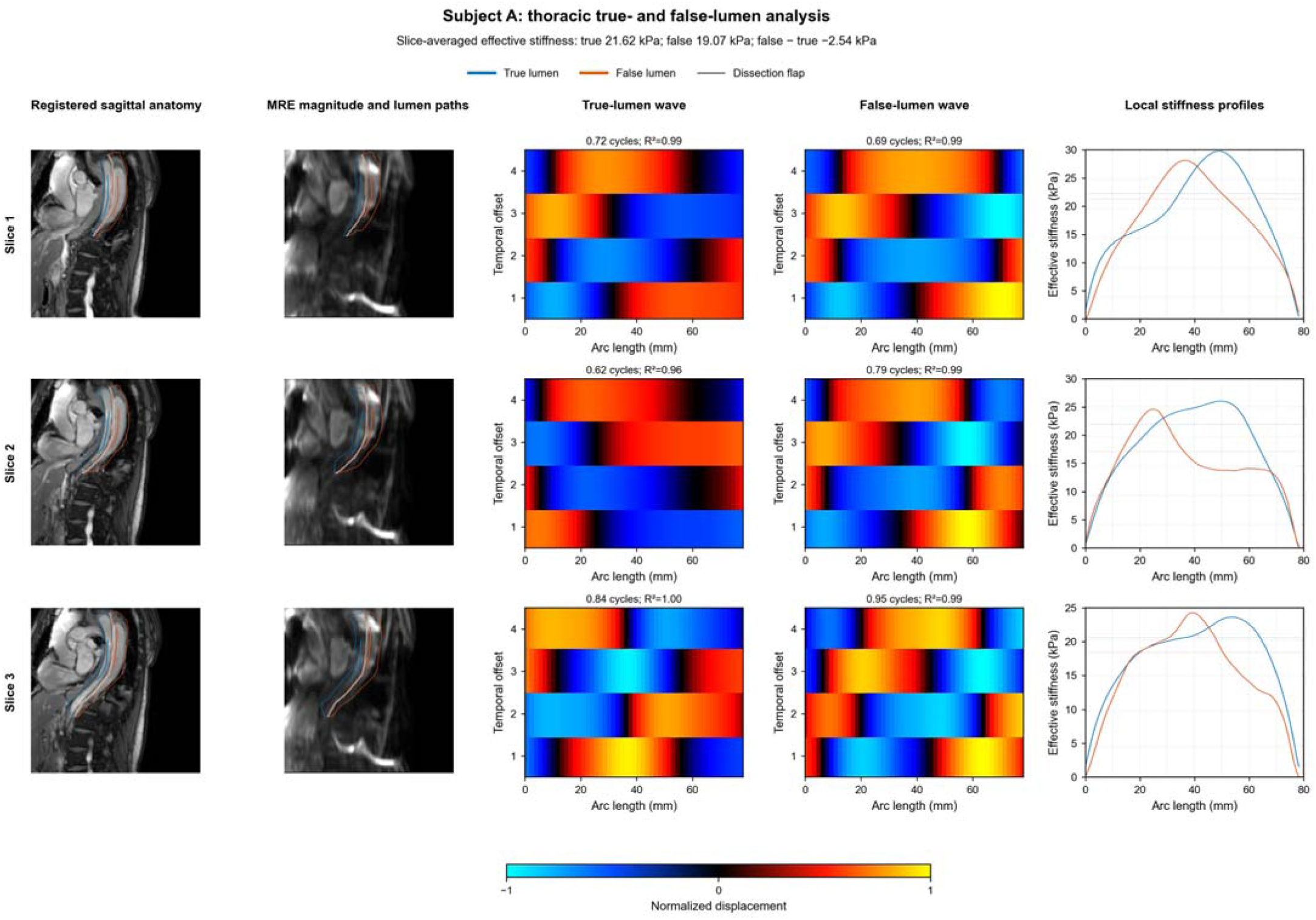
Complete Subject A thoracic true and false-lumen review. Sagittal anatomy, matched MRE magnitude and lumen paths, true- and false-lumen traveling-wave profiles, and local-frequency stiffness profiles are shown for all three analyzed slices. Wave color encodes signed displacement normalized within each displayed lumen. Dashed lines in the stiffness profiles indicate the corresponding central-path means. All six lumen-specific measurements passed the prespecified quality control criteria.

**Supplemental Figure 3.**
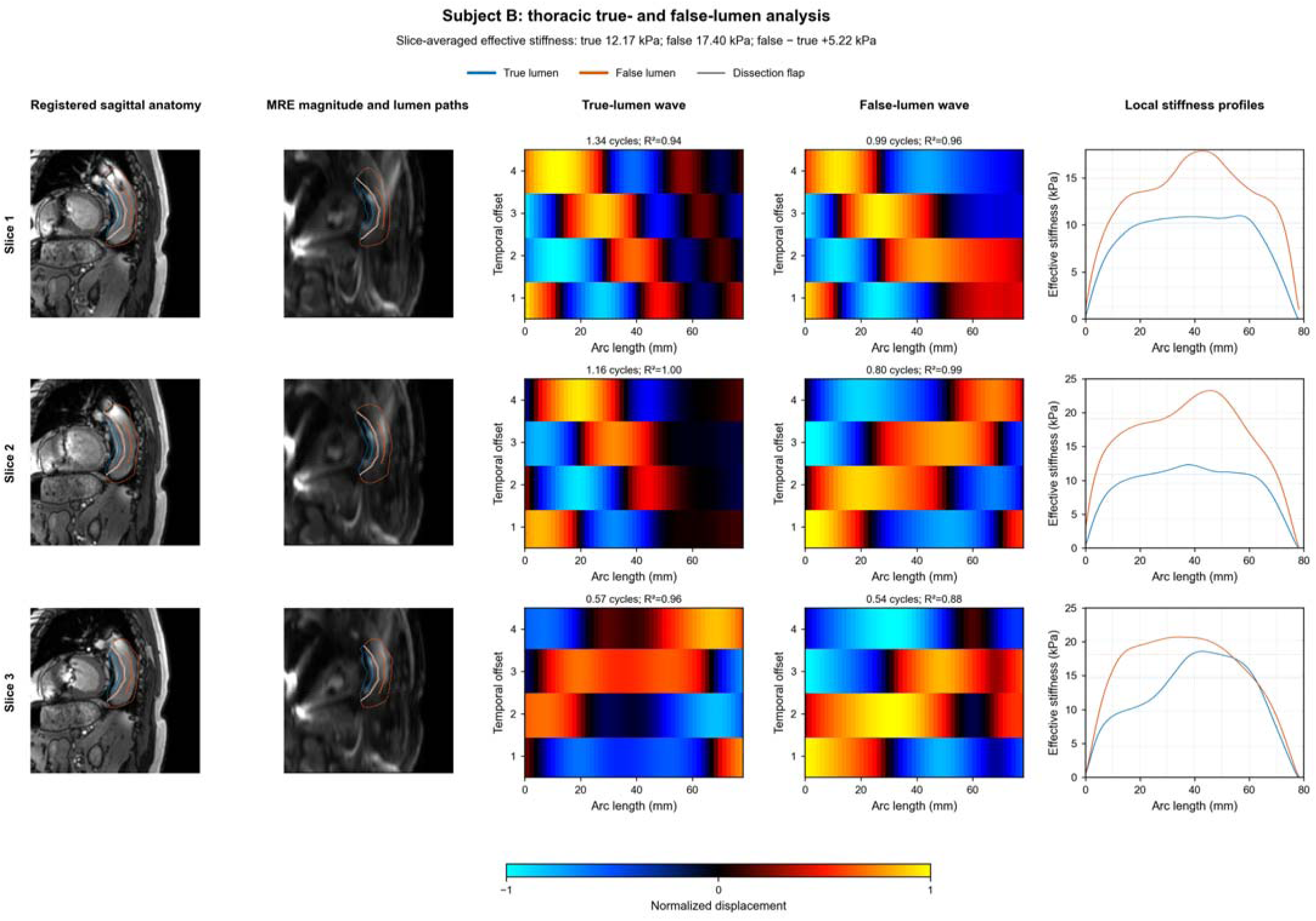
Complete Subject B thoracic true and false-lumen review. Sagittal anatomy, matched MRE magnitude and lumen paths, true- and false-lumen traveling-wave profiles, and local-frequency stiffness profiles are shown for all three analyzed slices. Wave color encodes signed displacement normalized within each displayed lumen. Dashed lines in the stiffness profiles indicate the corresponding central-path means. All lumen-specific profiles passed the prespecified quality control thresholds.

